# Lifetime Earnings in Pediatric Cardiology: A Net Present Value Analysis of Academic and Private Practice Pathways

**DOI:** 10.1101/2025.08.27.25334609

**Authors:** Murad Almasri, Lubaina Ehsan, Abdelrahman F. Masri, Mohammed Ayyad, Krittika Joshi, Joshua Daily

## Abstract

**Background:** Pediatric cardiology requires extensive training, yet the long-term financial implications across academic subspecialties and private practice remain poorly defined. Clearer understanding is essential for career decisions and workforce planning.

**Methods:** We used a net present value (NPV) framework to model lifetime earnings for pediatric cardiologists across three academic subspecialties (diagnostic, cardiac intensive care, interventional) under five promotion trajectories, comparing them with private practice. Compensation data came from the Association of Academic Administrators in Pediatrics, the Association of American Medical Colleges, and the Medical Group Management Association. Monte Carlo simulations and sensitivity analyses accounted for variation in salary percentile, discount rate, and career length.

**Results:** Lifetime earnings were substantial across all pediatric cardiology pathways, with NPVs exceeding $7 million(M) in nearly all scenarios. Interventional cardiology yielded the highest NPV at the 50th percentile under a typical academic promotion trajectory ($7.99M), followed by cardiac intensive care ($7.76M) and diagnostic cardiology ($7.00M). Private practice produced an NPV of $7.08M at the 50th percentile, with a ramp-up model increasing this to $7.30M; still below interventional and CICU academic tracks. Academic earnings increased by up to $2.44M through early promotion compared to no promotion, and by up to $867,000 through leadership roles, depending on subspecialty. Salary percentile was the most influential NPV driver; interventional cardiology at the 90th percentile exceeded $10.4M, and private practice reached $10.76 million. Private practice exhibited the widest lifetime earnings range.

**Conclusions:** Pediatric cardiologists, particularly those in interventional subspecialties or academic leadership, achieve substantial lifetime earnings. At the 50th percentile, academic and private practice careers offer comparable financial outcomes, but private practice shows greater variability. Optimizing academic pathways through early promotion, high-percentile salaries, or leadership roles can match or exceed private practice. These findings can guide trainees and inform institutional strategies for recruitment, and compensation equity in pediatric cardiology.

**What is Known:**

- Pediatric subspecialists, including cardiologists, have historically reported lower compensation than adult medicine counterparts.
- Lifetime earning disparities across the pediatric workforce are documented, but subspecialty-specific financial outcomes within pediatric cardiology remain uncharacterized.

**What the Study Adds:**

- Applies a net present value framework with Monte Carlo simulations to model lifetime earnings across academic pediatric cardiology subspecialties and private practice.
- Demonstrates that academic pediatric cardiology, especially interventional and CICU tracks, can achieve lifetime earnings comparable to or greater than private practice.
- Identifies promotion timing, leadership roles, and higher salary percentiles as major modifiable drivers of lifetime earnings, providing actionable insights for trainees and institutions.

## Introduction

Pediatric cardiology is one of the most rigorous and highly specialized fields within pediatrics (1, 2), requiring a prolonged training pathway that typically includes three years of pediatric residency, three years of cardiology fellowship, and often one or more years of advanced subspecialty training. Despite this intensive preparation, pediatric cardiologists, like many pediatric subspecialists, often face a financial disadvantage compared to their peers in adult medicine(3). Prior work has documented wide disparities in lifetime earnings across the pediatric workforce(4), but no studies have examined differences within pediatric cardiology itself.

Trainees considering a career in pediatric cardiology must weigh their passion for the field against concerns about educational debt, delayed earning potential, and a prolonged training timeline (5). These financial considerations are particularly relevant when deciding between academic and private practice settings, or when selecting among subspecialties such as interventional cardiology, cardiac intensive care, and imaging. In many areas of medicine, pursuing an academic career is often associated with accepting lower lifetime earnings in exchange for perceived benefits like research opportunities, teaching responsibilities, or institutional stability(6). However, it is unclear whether this trade-off holds true in pediatric cardiology, where compensation models, procedural intensity, and leadership roles may significantly influence long-term financial outcomes.

Despite the availability of national salary data from sources such as the Association of American Medical Colleges (AAMC), Medical Group Management Association (MGMA), and Association of Administrators in Academic Pediatrics (AAAP), there has been no systematic analysis comparing long-term financial outcomes across pediatric cardiology career paths.Trainees and mentors currently lack accessible models to evaluate how factors like promotion trajectory, leadership roles, and compensation percentile affect long-term earnings in the field.

Net Present Value (NPV)(7) analysis offers a robust and widely used financial framework to evaluate lifetime earnings by accounting for both the timing and amount of income. It is especially useful in medicine, where career trajectories span multiple decades, and financial decisions made early in training can have compounding effects over time. While NPV modeling has been applied to other specialties, it has not been previously used to evaluate pediatric cardiology in this level of detail.

In this study, we apply NPV analysis to compare lifetime earnings for pediatric cardiologists across three academic subspecialties (diagnostic, cardiac intensive care, and interventional) and private practice. Using real-world compensation data from AAAP, AAMC, and MGMA, we model five academic promotion trajectories and two private practice salary structures. To reflect real-world uncertainty in financial and career assumptions, we also conduct Monte Carlo(8) simulations and sensitivity analyses varying salary percentile, discount rate(9), and career duration. Our goal is to provide pediatric cardiology trainees, mentors, and institutional leaders with a comprehensive, data-driven framework to inform career decision-making and workforce planning.

## Methods

We used a NPV framework to model lifetime earnings for pediatric cardiologists across academic and private practice career paths. Compensation data were sourced from three national databases: AAAP for subspecialty-specific academic salaries, AAMC for all academic pediatric cardiology subspecialties, and the MGMA for private practice.

### Career Tracks and Promotion Scenarios

Three academic pediatric cardiology subspecialties; diagnostic, cardiac intensive care, and interventional; were modeled under five distinct promotion scenarios:

1. Typical Promotion: 7 years as Assistant Professor, 7 years as Associate Professor, remainder as Full Professor.
2. Early Promotion: 5 years as Assistant, 5 years as Associate, remainder as Full Professor.
3. Late Promotion: 10 years as Assistant, remainder as Associate Professor.
4. No Promotion: Entire career as Assistant Professor.
5. Leadership Track: 7/7/7 progression through academic ranks, followed by 12 years at the 90th percentile salary for division chiefs or section heads.

Academic pediatric cardiology (all subspecialties) was modeled using AAMC salary data, assuming the typical promotion trajectory.

### Private Practice Models

Private practice was modeled using MGMA data under two frameworks:

- Fixed Model: Salary held constant at a selected percentile (25th, 50th, 75th, or 90th) throughout the career.
- Ramp-Up Model: Simulated progressive salary growth—starting at the 25th percentile for years 1–3, the 50th percentile for years 4–10, the 75th percentile for years 11–20, and returning to the 50th percentile for years 21–32.

### Base Assumptions

We assumed physicians began their attending careers at age 35 following medical school, residency, cardiology fellowship, and one additional year of advanced training. All base-case scenarios modeled a 32-year clinical career, ending at age 67, which corresponds to the full retirement age defined by the U.S. Social Security Administration. We also included alternate models to reflect career variability:

- Shortened Career: Retirement at age 62 (27 years)
- Extended Career: Retirement at age 72 (37 years)

### NPV Calculations

NPV was calculated using the standard financial formula:

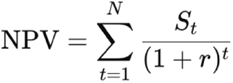

Where:

- St= salary in year t
- *r* = real discount rate (base case: 3%)
- *N* = career duration (in years)

All earnings were expressed in constant 2024 U.S. dollars, assuming no real salary growth over time.

### Monte Carlo Simulation

To reflect real-world variability in economic conditions and career trajectories, we performed a 10,000-iteration Monte Carlo simulation for each major career path. The following parameters were randomly varied in each iteration:

- Discount rate: Uniform distribution from 2% to 5%
- Career duration: Triangular distribution with minimum 27, mode 32, and maximum 37 years
- Salary percentile: Randomly selected from the 10th, 25th, 50th, 75th, or 90th percentile

Academic careers followed a typical promotion schedule, while private practice salaries were modeled as fixed at the selected percentile. The resulting simulations were used to generate violin plots and histograms representing the distribution of lifetime NPVs.

### Sensitivity Analyses

We conducted one-way sensitivity analyses to isolate the effects of discount rate and career length. NPVs were recalculated under the following variations:

- Discount rate: 0%, 3%, and 5%
- Career duration: 27, 32, and 37 years

## Results

All pediatric cardiology career pathways evaluated yielded substantial lifetime earnings, with NPVs exceeding $7 million across nearly all scenarios (Figure 1). Under the base-case model; a 32-year career, typical academic promotion trajectory, and 50th salary percentile; interventional cardiology produced the highest NPV at $7.99 million. This was followed by cardiac intensive care at $7.76 million and diagnostic cardiology at $7.00 million. The AAMC-based model, representing a pooled combination of all academic pediatric cardiology subspecialties, yielded an NPV of $7.43 million, closely aligning with the subspecialty-specific estimates from the AAAP. This consistency between independent salary data sources supports the reliability of these modeled outcomes.

**Figure 1:**
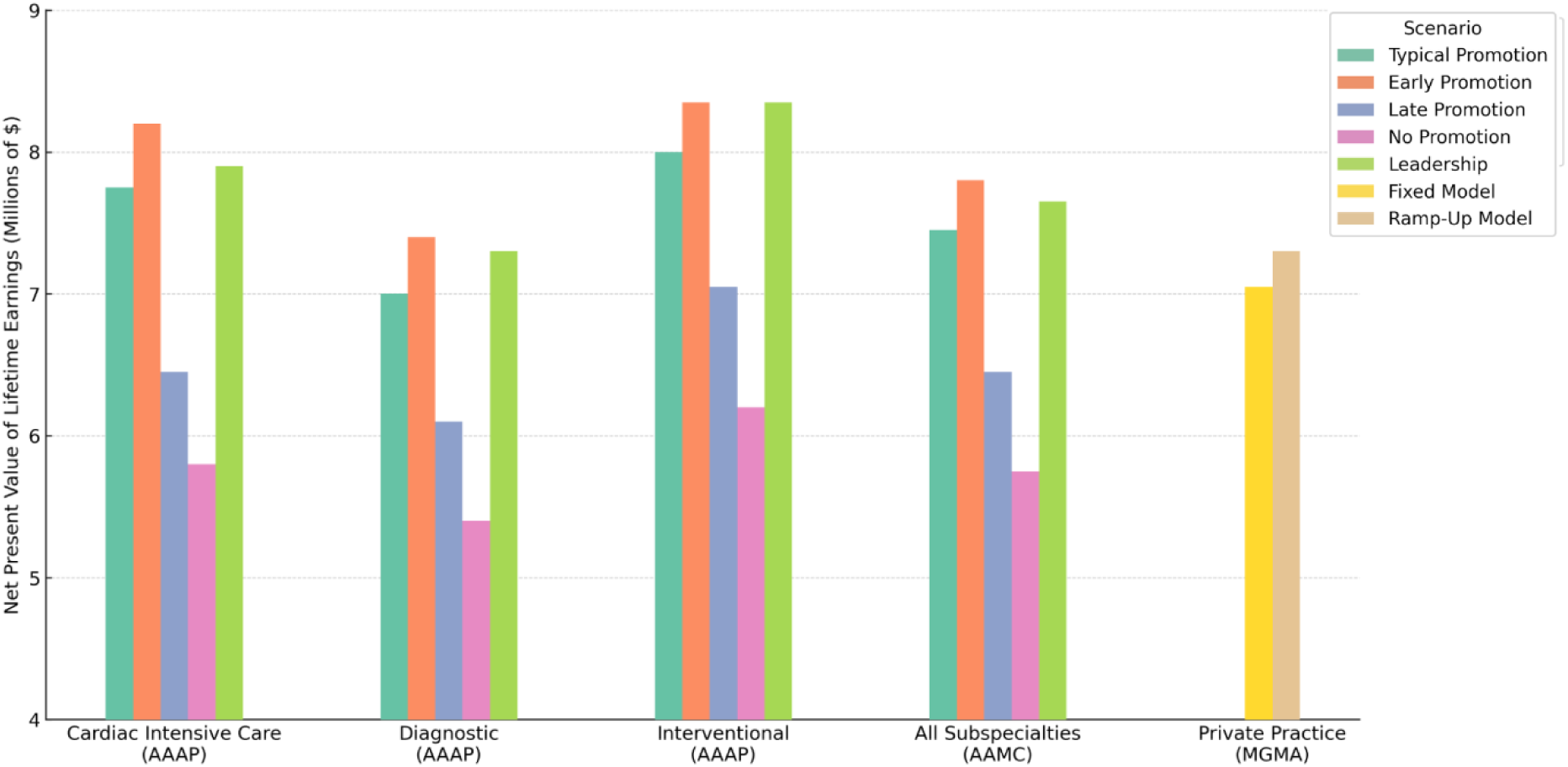
Lifetime Earnings by Pediatric Cardiology Career Track. This chart shows the net present value (NPV) of lifetime earnings across five career paths: cardiac intensive care, diagnostic, interventional, academic-general, and private practice. Academic tracks are modeled under five promotion scenarios. Private practice includes both fixed and ramp-up salary models. NPVs assume a 32-year career and are reported in millions of 2024 USD.

In comparison, private practice pediatric cardiology yielded an NPV of $7.08 million at the 50th percentile using a fixed salary model. A more optimistic ramp-up model, which simulates progressive salary increases over the first 20 years, produced an NPV of $7.30 million. Notably, even diagnostic academic cardiologists, who represent the lowest-earning academic subspecialty in our model, achieved lifetime earnings nearly equivalent to the private practice fixed model. These findings suggest that academic pediatric cardiology - across all subspecialties - can be financially competitive with private practice, particularly when factoring in promotion and leadership opportunities.

At higher salary percentiles, the gap narrows or reverses. As shown in Figure 2, private practice at the 90th percentile yielded an NPV of $10.76 million, narrowly surpassing interventional cardiology, which reached $10.42 million. Cardiac intensive care followed at $9.69 million, and diagnostic cardiology at $8.56 million. However, at lower percentiles, academic interventional and CICU pathways consistently outperformed private practice, emphasizing the importance of both subspecialty and compensation level in determining financial outcomes.

**Figure 2:**
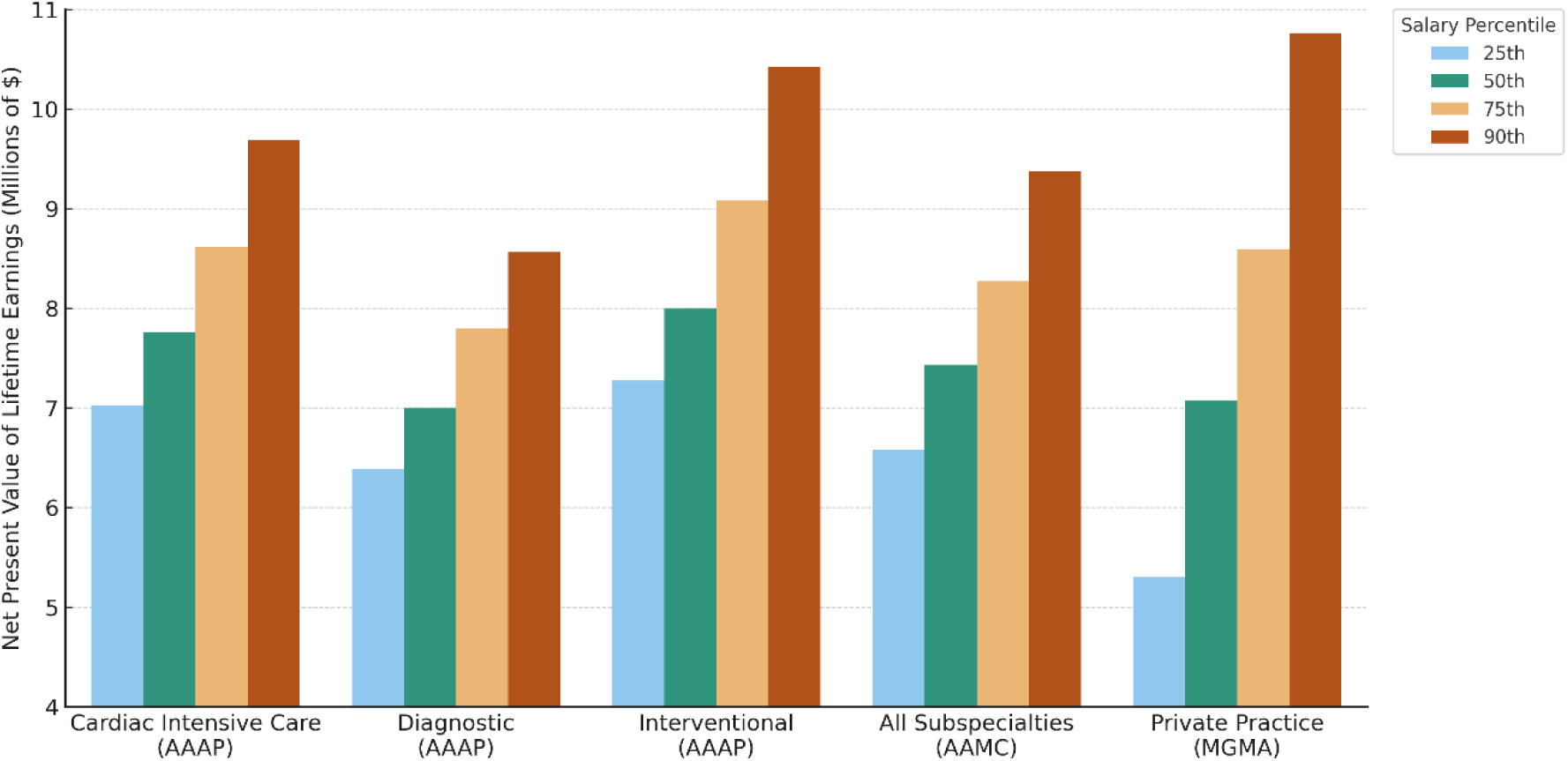
Earnings by Salary Percentile Across Career Paths. NPVs are displayed for each career path at the 25th, 50th, 75th, and 90th salary percentiles. Academic models use a typical promotion timeline; private practice assumes fixed salaries. Higher percentiles yield greater earnings, especially for interventional cardiology and private practice.

Figure 3 highlights the impact of career length. Extending the career from 27 to 37 years increased NPVs by more than $3 million across most paths. For example, interventional cardiology grew from $6.78 million to $9.98 million. The rank order of subspecialties remained stable across all durations.

**Figure 3:**
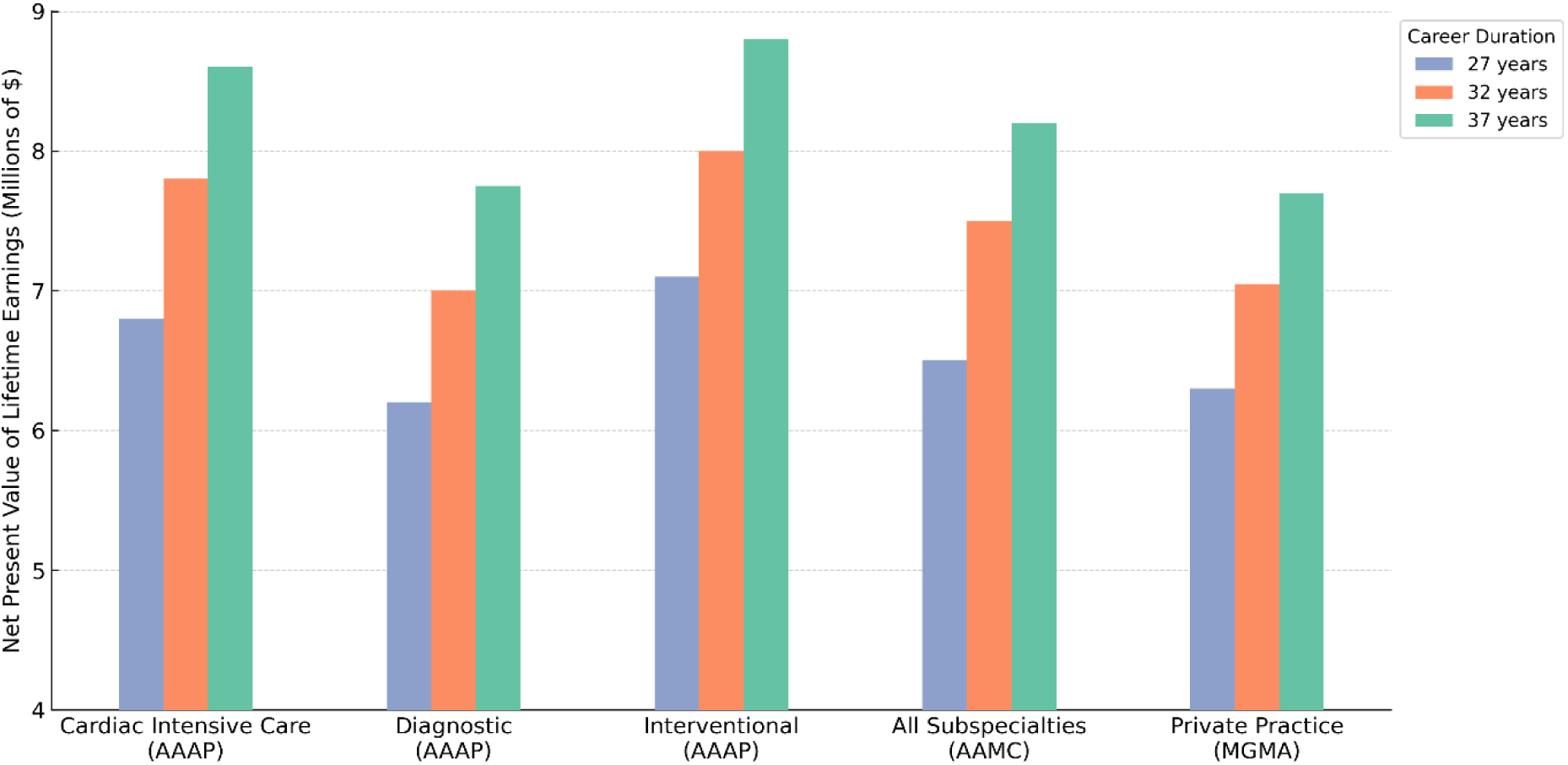
Impact of Career Length on Lifetime Earnings. This figure compares NPVs across three career durations: 27, 32, and 37 years. All values are shown in millions of 2024 USD. Longer careers significantly increase earnings, but the ranking of career tracks remains stable.

Promotion trajectory and leadership roles were powerful modifiers of long-term earnings. As illustrated in Figure 4, moving into academic leadership increased NPVs by up to $1.5 million. At the 90th percentile, a leadership-track interventional cardiologist earned $10.82 million, compared to $8.56 million for a peer following a typical promotion pathway. In contrast, remaining at the assistant professor level yielded substantially lower NPVs, often trailing even the 25th percentile of private practice.

**Figure 4:**
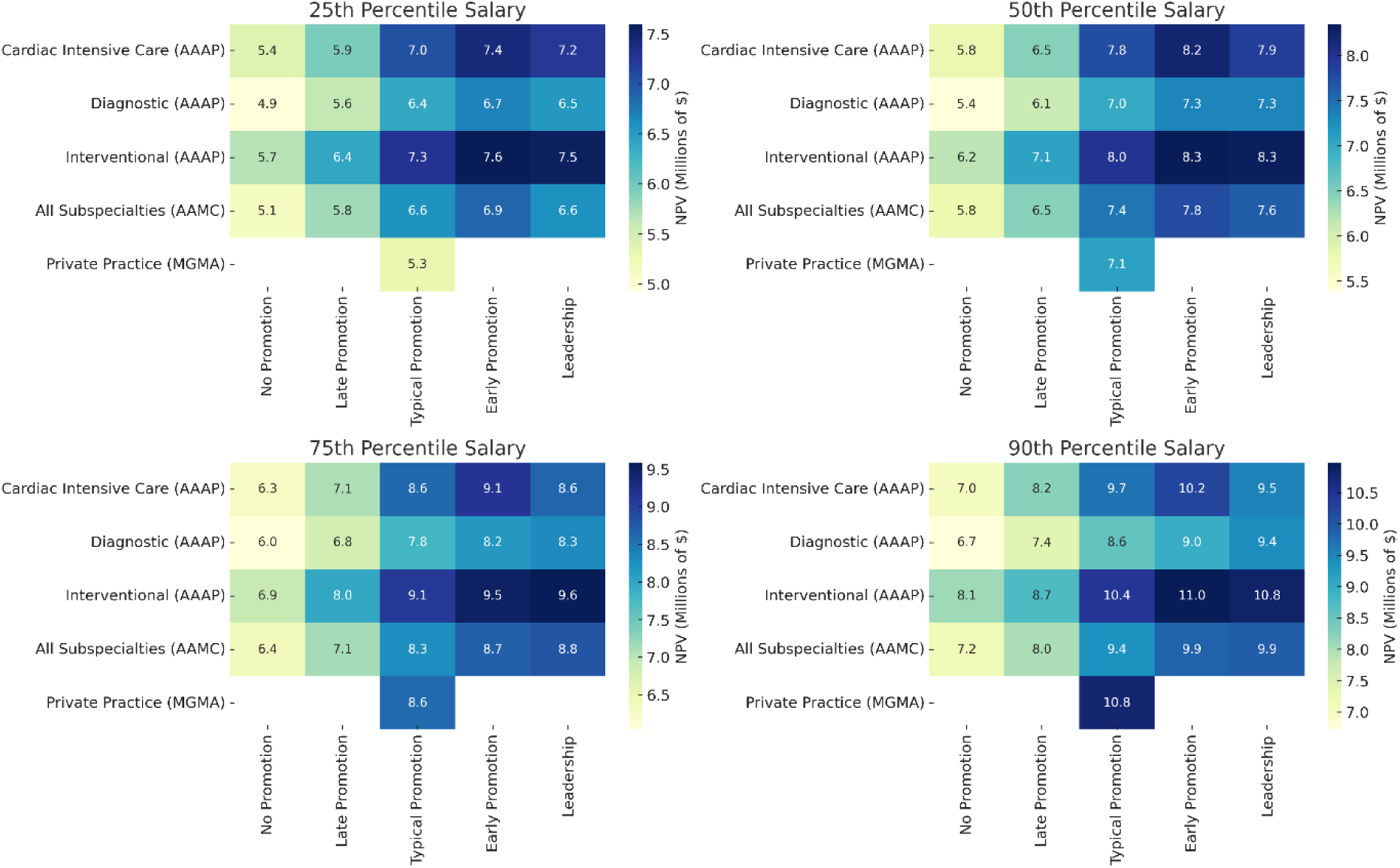
Combined Effects of Promotion and Salary Percentile. Heatmaps illustrate how promotion timing and salary percentile affect NPVs. Leadership roles and early promotion increase earnings, especially in interventional and CICU pathways. No promotion scenarios yield the lowest NPVs.

The Monte Carlo simulation results (Figure 5) demonstrated that interventional cardiology and private practice had the highest median NPVs and greatest upside variability. However, academic tracks with favorable promotion timelines or leadership roles showed competitive or superior outcomes in many simulations. Academic pathways also exhibited narrower interquartile ranges, suggesting more predictable earnings over time.

**Figure 5:**
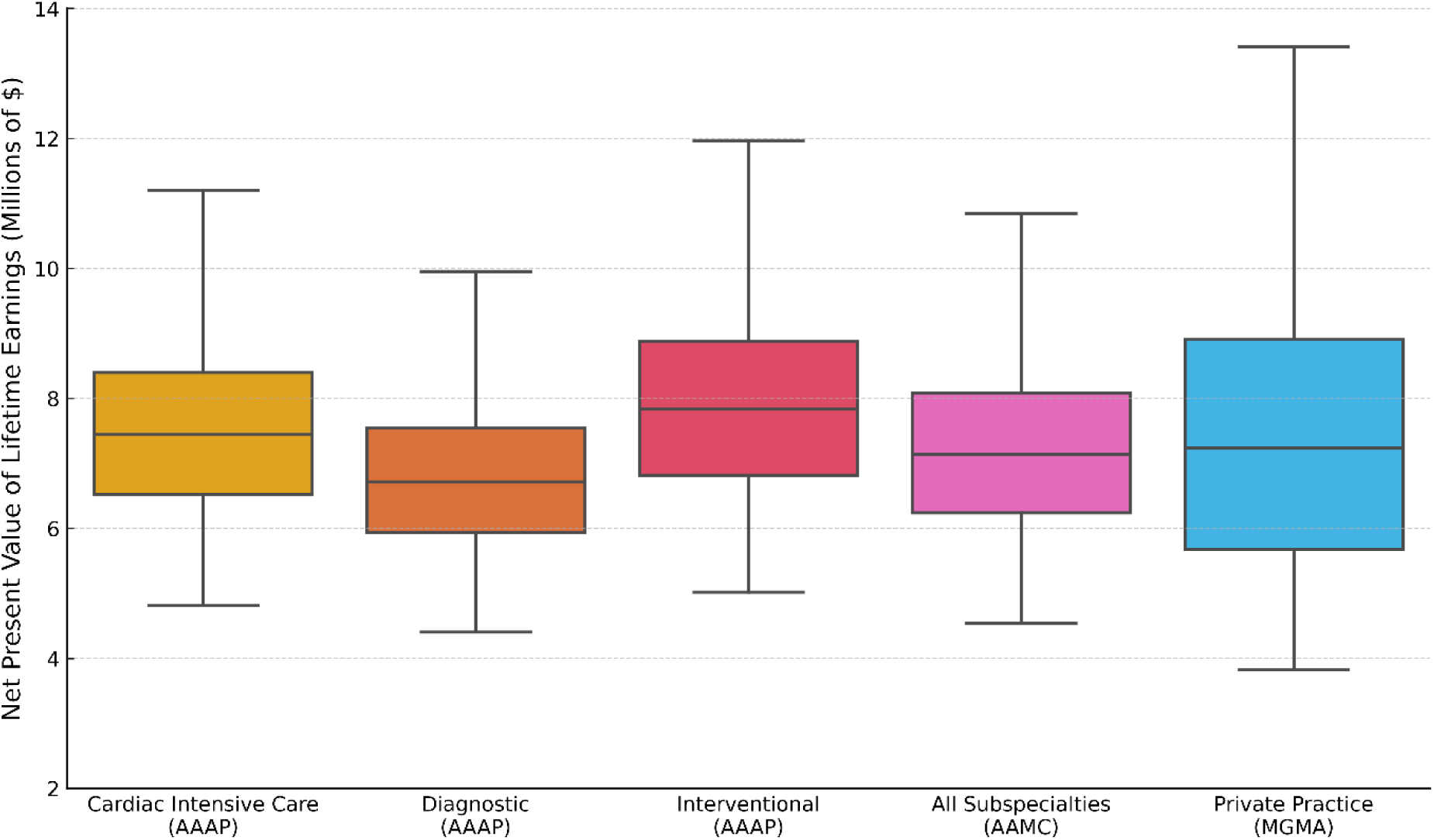
Monte Carlo Simulation of Earnings. This simulation models 10,000 potential lifetime earning scenarios, incorporating variation in salary, discount rate, and career length. Interventional cardiology and private practice show the greatest upside and variability.

Finally, sensitivity analysis (Figure 6) confirmed that discount rate, salary percentile, and career length were the most influential variables affecting NPV. As expected, higher discount rates reduced overall NPVs but did not meaningfully alter the relative ranking of career paths. Interventional cardiology remained the most financially resilient pathway across all modeled scenarios.

**Figure 6:**
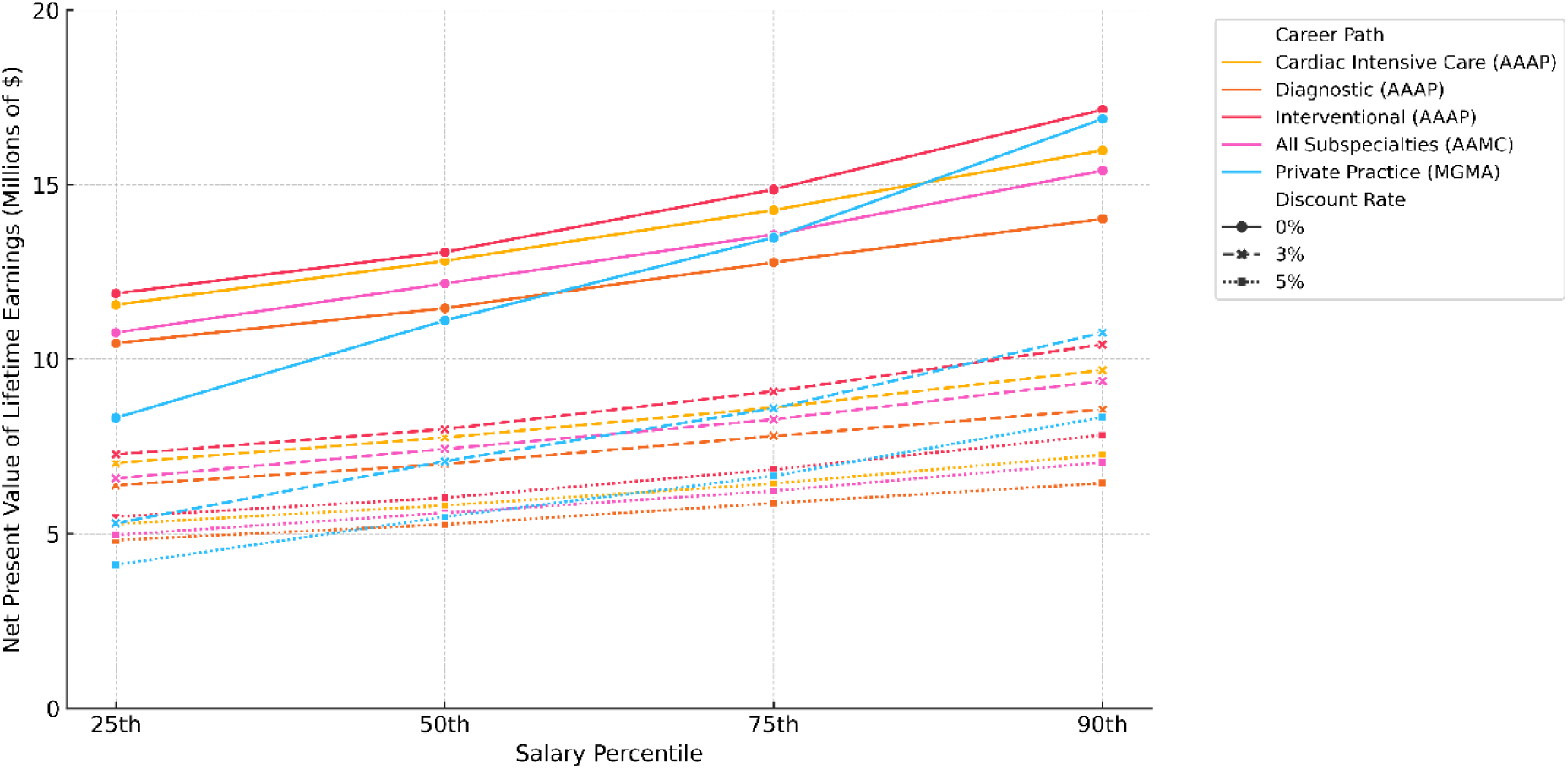
Sensitivity of Earnings to Financial Assumptions. This chart explores how NPVs shift with changes in discount rate and salary level. Despite variability, interventional cardiology consistently offers the highest projected earnings.

## Discussion

This is the first study to apply NPV modeling to evaluate lifetime earnings across pediatric cardiology subspecialties and career tracks. By incorporating real-world compensation data, academic promotion timelines, leadership roles, and Monte Carlo simulations, we provide a comprehensive and nuanced financial analysis of pediatric cardiology career pathways. Our findings reveal three main insights: first, despite the extensive training required, pediatric cardiologists achieve substantial lifetime earnings; placing them among the highest earners within pediatrics. Second, academic pediatric cardiologists earn lifetime incomes that are comparable to those in private practice, challenging long-held assumptions seen in other medical specialties(6). Third, lifetime earnings in academic settings can be significantly increased through strategic decisions such as selecting higher-paying subspecialties, attaining early promotion, and negotiating for higher salary percentiles.

First, despite long training durations, all pediatric cardiology career tracks evaluated in this study yielded strong financial outcomes. Every modeled pathway; including those in academic and private settings; produced net present values exceeding $7 million over a 32- year career. At the 50th percentile, interventional academic cardiologists earned $7.99 million, cardiac intensivists earned $7.76 million, and diagnostic cardiologists earned $7.00 million. These values place pediatric cardiologists among the highest earners within pediatrics and are consistent with findings from national surveys that rank pediatric cardiology among the top-compensated pediatric subspecialties(4). However, it is important to acknowledge that pediatric cardiologists still earn substantially less than their adult cardiology counterparts(3). In one national analysis, pediatric cardiologists had nearly $2 million lower lifetime earnings compared to adult cardiologists(4). While prior work has focused on the pediatric–adult earnings gap, our findings emphasize that within pediatrics, cardiology; regardless of subspecialty; offers compelling long-term earning potential. This insight may be particularly important to trainees weighing the opportunity cost of prolonged training.

Second, our analysis challenges the assumption that choosing an academic career requires a substantial financial sacrifice(10)(11). At the 50th percentile, diagnostic academic cardiologists earned nearly the same lifetime NPV as private practice physicians ($7.00 million vs. $7.08 million, respectively), while interventional and CICU academic cardiologists outperformed private practice. Even the AAMC-based model, which reflects pooled academic subspecialty data, produced an NPV of $7.43 million, closely aligning with subspecialty-specific estimates from AAAP. Notably, private practice exhibited the widest distribution of lifetime earnings, with high variability depending on compensation percentile. At the 90th percentile, private practice NPV rose to $10.76 million, exceeding all academic scenarios except leadership-track interventional cardiology, while at lower percentiles, private practice earnings were outpaced by many academic pathways. These findings contrast with literature from adult medicine, where academic physicians often earn significantly less than their private practice counterparts over a career(6)(12). In pediatric cardiology, academic roles, particularly in procedural subspecialties or leadership positions, can offer earnings that rival or exceed private practice. This trend may reflect a narrower salary gap in pediatrics and a more standardized academic compensation structure across institutions Third, academic career earnings are highly modifiable and can be substantially improved through strategic decisions. Among all variables analyzed, salary percentile was the most influential driver of lifetime NPV. Moving from the 25th to the 90th percentile increased earnings by $3.15 million for interventional cardiologists, $2.66 million for CICU physicians, and $2.17 million for diagnostic cardiologists. Promotion trajectory also had a substantial impact: avoiding delayed promotion increased NPV by $910,000 to $1.31 million, while avoiding no promotion increased earnings by $1.61 million to $1.99 million, depending on subspecialty. Subspecialty choice contributed a smaller, but still meaningful, difference of approximately $1.00 million between the highest- and lowest-earning academic tracks at the 50th percentile. While salary percentile is influenced by institutional policies, it is often closely tied to clinical productivity and RVU generation(13). Pediatric cardiologists should understand how their compensation is determined; in many cases, producing more RVUs or exceeding productivity benchmarks is key to achieving a higher salary percentile. These findings highlight that clinical performance, salary negotiation, and promotion timing are just as important as subspecialty choice. Institutions that provide transparent compensation structures and equitable access to promotion and leadership can help support academic retention(14). Likewise, trainees and junior faculty may benefit from mentorship on career advancement and negotiation as essential components of long-term financial planning(15).

This study has several limitations. First, while we used nationally available compensation data from the AAAP, AAMC, and MGMA, actual salaries can vary by institution, geography, and individual negotiation. We modeled standard promotion and leadership timelines, but career paths are inherently variable and can include part-time work, administrative responsibilities, or changes in clinical scope. Second, although our ramp-up model reflected private practice salary progression, we assumed static promotion-based salaries for academic careers and did not model year-over-year salary increases. Third, we did not account for debt burden, benefits, cost of living, or tax effects, all of which may meaningfully impact real-world financial outcomes. Finally, our analysis focuses solely on financial metrics. Non-financial considerations, such as academic mission, research opportunities, job satisfaction, mentorship, and lifestyle, are critical to career decision-making but fall outside the scope of this economic model.

Pediatric cardiology offers substantial lifetime earnings across both academic and private practice pathways. Contrary to assumptions drawn from adult specialties, academic pediatric cardiologists, particularly those in procedural subspecialties or leadership roles, can achieve lifetime incomes comparable to or exceeding those in private practice. Within academic medicine, earnings are highly modifiable: salary percentile, promotion timing, and subspecialty choice each meaningfully affect long-term financial outcomes. These data provide a practical framework to inform career planning for trainees and support institutional strategies aimed at equitable compensation, faculty retention, and leadership development in pediatric cardiology.

## Data Availability

Data can be shared upon request.

## Acknowledgments

None

## Funding

None

## Conflicts of Interest

The authors declare no conflicts of interest.

## Notes

### Competing Interest Statement

The authors have declared no competing interest.

### Funding Statement

No funding was received.

### Author Declarations

The study was exempted from IRB (no patient identifiers).

